# COVID-19 containment policies through time may cost more lives at metapopulation level

**DOI:** 10.1101/2020.04.22.20075093

**Authors:** Konstans Wells, Miguel Lurgi

## Abstract

The rapid and pandemic spread of COVID-19 has led to unprecedented containment policies in response to overloaded health care systems. Disease mitigation strategies require informed decision-making to ensure a balance between the protection of the vulnerable from disease and the maintenance of global economies. We show that temporally restricted containment efforts, that have the potential to flatten epidemic curves, can result in wider disease spread and larger epidemic sizes in metapopulations. Longer-term rewiring of metapopulation networks or the enforcement of feasible long-term measures that decrease disease transmissions appear to be more efficient than temporarily restricted intensive mitigation strategies (e.g. short-term mass quarantine). Our results may inform balanced containment strategies for short-term disease spread mitigation in response to overloaded health care systems and longer-term epidemiological sizes.

## Main Text

The rapid and unprecedented pandemic spread of the newly emerged coronavirus SARS-CoV-2 imposes a significant burden on national health systems, with alarmingly high fatality and hospitalization rates (*1*). While this novel virus has affected the human population worldwide, severe cases are mostly confined to elderly people above 70 years old, and those with underlying conditions such as cardiovascular and respiratory disease, cancer, or diabetes (*2*). Immediate national responses have been numerous and varied. Nonetheless, by early April 2020, only four months after the first records of the virus in Wuhan, China, a considerable proportion of the world population have found themselves being mass-quarantined through ‘stay-at-home’ policies.

Without any protective pharmaceutical treatment (e.g. vaccination) available, disease control is a challenging task because it relies solely on combating pathogen spread resulting from the myriads of host interactions. There is consensus that ‘flattening the epidemic curve’ by delaying disease spread through physical distancing measures, can lower the pressure on health services during the most critical periods of epidemic spread (*3, 4*). Nevertheless, the long-term and large-scale outcomes of intensive short-term mitigation strategies are little understood. Escaping infection at the individual level throughout the course of entire epidemics can only be achieved by permanently avoiding contacts with infectious agents. Hence, unless the disease is extirpated at the population level, the individual risk of infection can only be lowered by rigorous isolation or by depriving pool of susceptible individuals, hence lowering the force of infection through herd immunity (*5*).

The key assumption of homogeneous mixing of populations in traditional epidemiological models, which have played a central role in the study of the epidemic spread of COVID-19 so far (*3, 6*) are not met at large landscape scales. This is mainly due to the large heterogeneity in metapopulation networks and the spatial aggregation of individuals into different communities from household to city levels (*7*). In fact, it is well known that contact network structure and patterns of connectivity among local populations are main drivers of contagion processes from local to regional scale, with contact frequencies among populations largely determining whether local epidemics are coupled or not (*8*). However, the extent to which temporarily restricted intensive mitigation strategies (e.g. short-term total containment) in response to the COVID-19 epidemic can successfully lower overall epidemic size, and protect those most vulnerable, has yet to be quantified. and compared across different scenarios of landscape-level connectivity (i.e. metapopulation structure).

Theory predicts that metapopulation extinction-colonization dynamics can maintain disease persistence even with frequent local fade-out and extirpation (*9*). If local epidemics in different populations are weakly coupled, synchronous but short-term mitigation strategies may fail their target to contain disease spread during epidemic peaks (*10*). Further, local containment of outbreaks with delayed local extirpation may facilitate disease spread at the metapopulation level. Metapopulation source-sink dynamics, the effect of metapopulation structure, and the role of depleted pools of susceptible individuals (i.e. herd immunity), have been studied in the context of disease spread for decades (*11*). Nevertheless, there is a lack of understanding of whether and how temporally restricted intensive mitigation strategies, could result in long-term reduction of overall infection rates on spatially structured metapopulations. Understanding the usefulness of short-term intensive containment strategies constitutes a pressing challenge in times of the ongoing COVID-19 crisis. More generally, this understanding will contribute to the unresolved debate on whether it is possible to establish alternatives to socioeconomically detrimental mass quarantines (*6, 12*).

We address this challenge using an individual-based epidemiological model, that incorporates landscape-scale connectivity structure among local populations (i.e. metapopulation), to explore the outcome of the spread of COVID-19 and equivalent emerging infectious diseases in metapopulation networks after one year and under different mitigation scenarios. The interplay between transmission rates at local scales, and metapopulation dynamics (e.g. source-sink), which ultimately results in the possible depletion or replenishment of the pool of susceptible individuals, should determine the outcome of temporal mitigation strategies – either through escape from epidemic waves by accurately timed isolation or through more prolonged exposure induced by temporarily arrested and sustained epidemic spread.

### Modelling metapopulation epidemics with short-term containment efforts

We were interested in exploring the extent to which network structure and mitigation strategies may impact the outcomes of disease spread. We created synthetic metapopulations by linking discrete local populations according to three well known connectivity structures: random, scale-free and small world networks (*13*). We then modelled temporary mitigation on metapopulations as reduced transmission rates (the likely outcome of physical distancing measures) within certain time windows for the entire metapopulation. We additionally explored the effects of more selective mitigation strategies, in which transmission rates are only reduced for subsets of individuals assumed to be at high risk of severe disease effects (see methods).

On each of the three metapopulation structures, we ran numerical simulations of an individual-based SEIR model (see methods) across a sensible range of parameter values of (i) transmission rates (*β*), (ii) network connectivity (*C*), and individual commuter travel to (iii) neighbouring or (iv) distant populations (*δ* and *ρ*, respectively). We explored scenarios of lowering transmission rates (*ϕ*) between 10–90% over time windows of 21–300 days (*η*), which we believe to represent a range of possible mitigation strategies to prevent COVID-19 spread. This allowed us to compare the outcomes of different mitigation strategies in terms of relative epidemic sizes (the difference in epidemic sizes resulting from different mitigation strategies applied over the same transmission scenario) and spatial spread for different network types and mitigation strategies: (1) no mitigation, (2) whole-population mitigation, and (3) selective mitigation applied to vulnerable individuals only.

### Rewiring contact structures and constantly low transmission rates minimizes epidemics

Across all scenarios, we found strong support for scale-free connectivity structures among local populations being likely to minimize disease spread. Under no mitigation, total epidemic sizes were limited to <66% of the metapopulation infected and <30% local populations affected in scale-free networks (Fig. 1). Epidemic sizes and fractions of affected populations varied greatly among different transmission scenarios in random and small world networks, ranging from rapid disease extirpation with small epidemic sizes to spread into entire metapopulations (Fig. 1). This variation is mostly explained by differences in transmission rates *β* for all network types and connectivity *C* in scale-free networks (Table S2).

**Fig. 1.**
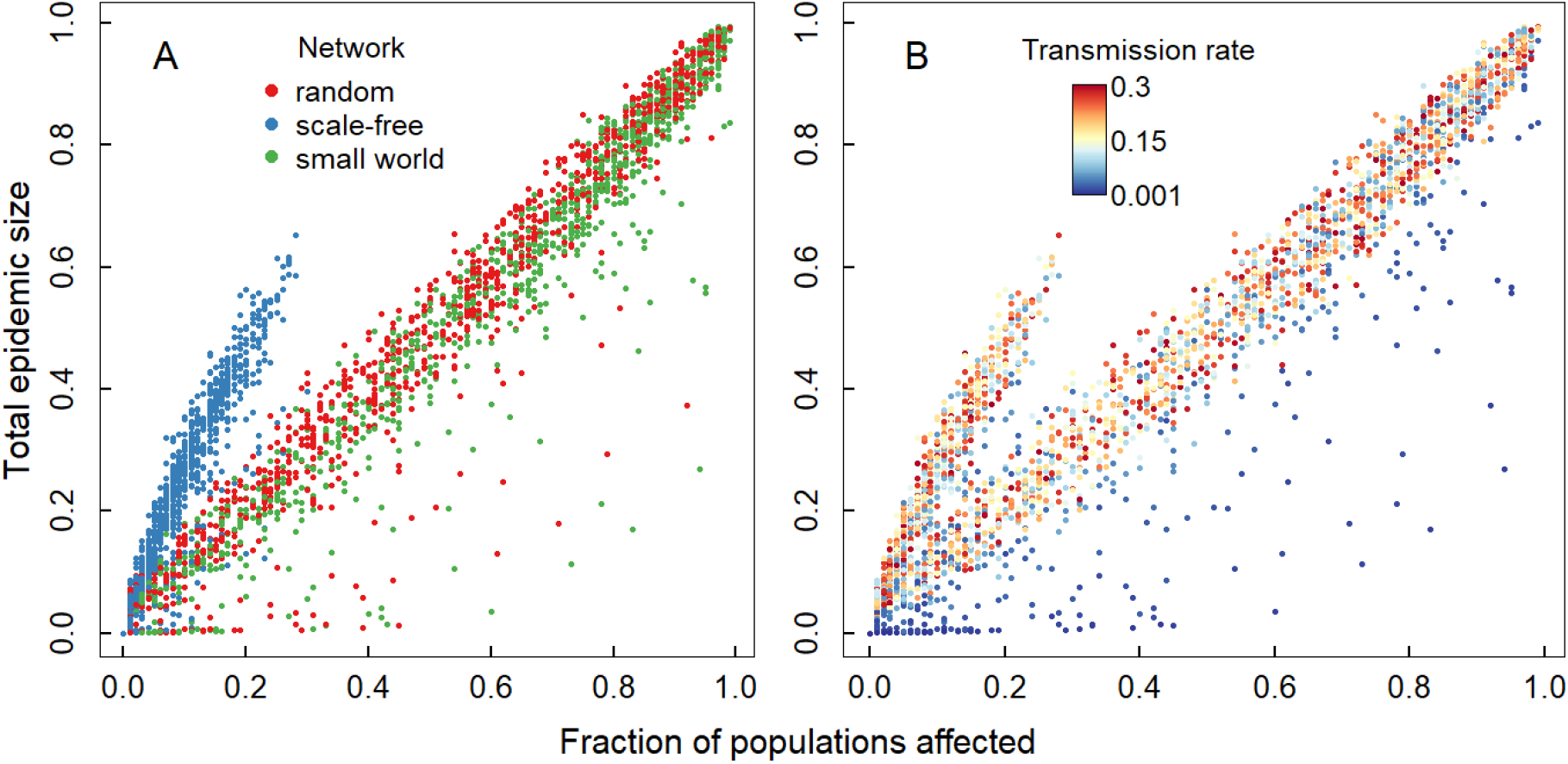
Fraction of populations affected and total epidemic size of COVID-19 metapopulation spread with different transmission scenarios. (A) Outcomes of disease spread with variable transmission rate, network type and connectivity, and commuter travel rate to neighbouring and distant populations coloured according to underlying network type. (B) Transmission scenarios as in (A) coloured according to the underlying transmission rates *β*.

### Short-term mitigation efforts are not always beneficial

While constantly low transmission rates (and hence low basic reproductive numbers R0) are the most crucial aspect in containing epidemic sizes and disease spread, we found rather intriguing and counterintuitive outcomes when temporary mitigation strategies are enforced. Epidemic size was smaller *without* any mitigation strategies in 29% of all transmission scenarios compared to scenarios where whole-population or selective mitigation strategies are enforced. In particular, no mitigation was likely to results in smaller relative epidemic sizes for transmission scenarios with large transmission rates *β* and weak containment efforts *ϕ* (Table S3). In contrast, relative epidemic size was smallest in 41% of transmission scenarios with whole-populations mitigation policy, especially for transmission scenarios with small *β*, high commuter travel *δ* and strong containment efforts *ϕ* (Table S3). Selective mitigation strategies, in turn, resulted in the smallest relative epidemic sizes and the lowest numbers of vulnerable individuals infected in transmission scenarios with large *β* in combination with weak containment efforts *η* and in scale-free networks (Table S3).

Notably, the fraction of populations affected increased significantly, with relative larger epidemic sizes, when mitigation strategies are implemented, compared to scenarios without mitigation, regardless of transmission rates (Fig. 2, Table S4). This indicates that mitigation policies may come at the cost of larger epidemic sizes if they do not contain disease within affected populations.

**Fig. 2.**
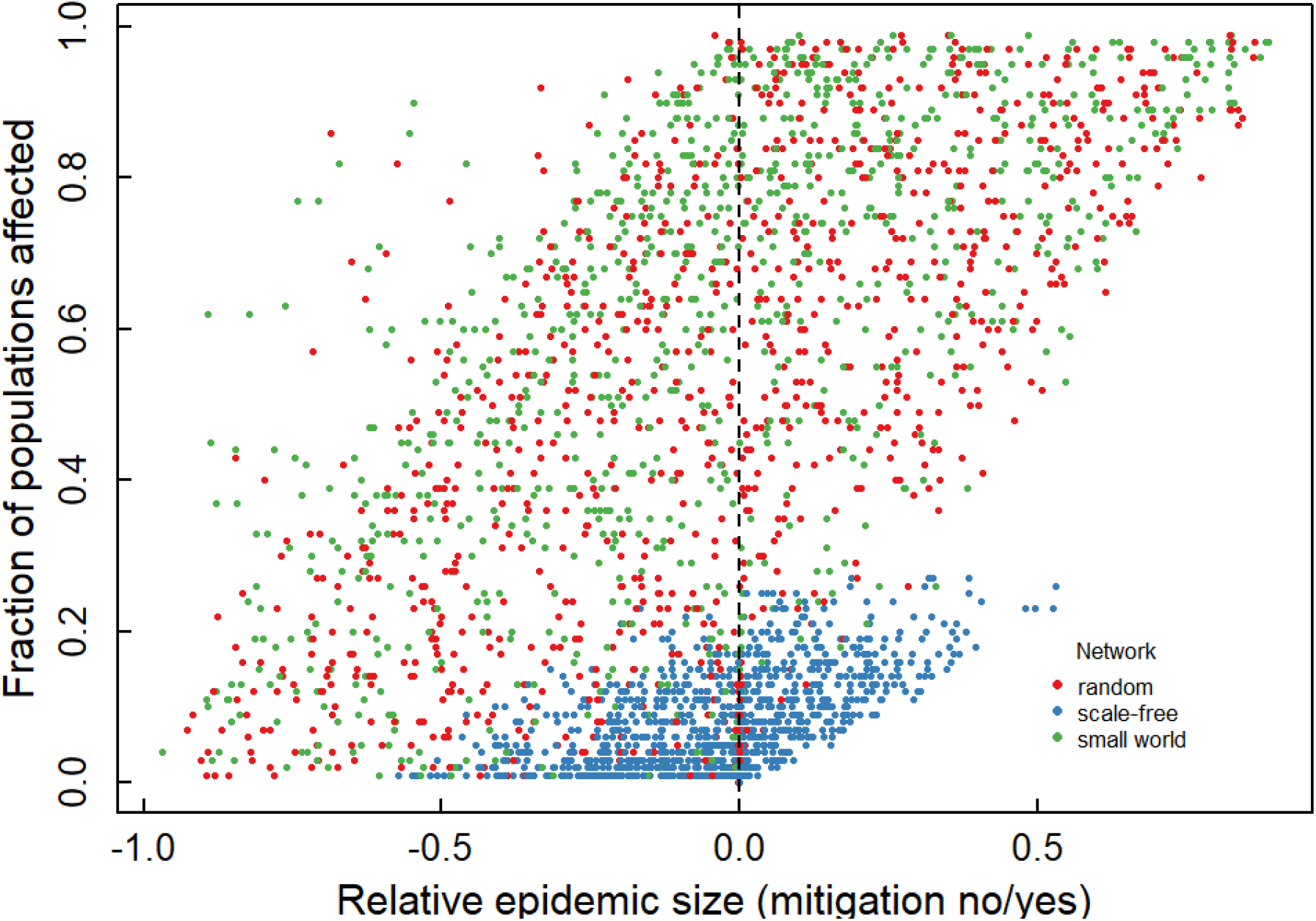
Relationship between metapopulation spread of COVID-19 and the relative difference in total epidemic sizes with temporary mitigation policy. Values >0 on the y-axis refer to larger epidemic sizes *with* short-term whole-population mitigation policies in place compared to the same transmission scenarios without any mitigation, whereas values <0 refer to transmission scenarios with larger epidemic sizes *without* mitigation policy. Colours depict different underlying network topographies.

As expected, epidemic peak size (relative to total epidemic size) was lowered with whole-population mitigation strategies, while increasing considerably for larger transmission rates *β* (Fig. 3, Table S5). There was however no apparent advantage of whole-population versus selective mitigation on epidemic peak sizes for vulnerable individuals only (Table S5). This highlights the crucial roles of transmission rates in driving the extents of epidemic peak sizes. Low transmission rates are thus key, along with strong short-term containment efforts, to effectively avoid overwhelming health care systems.

**Fig. 3.**
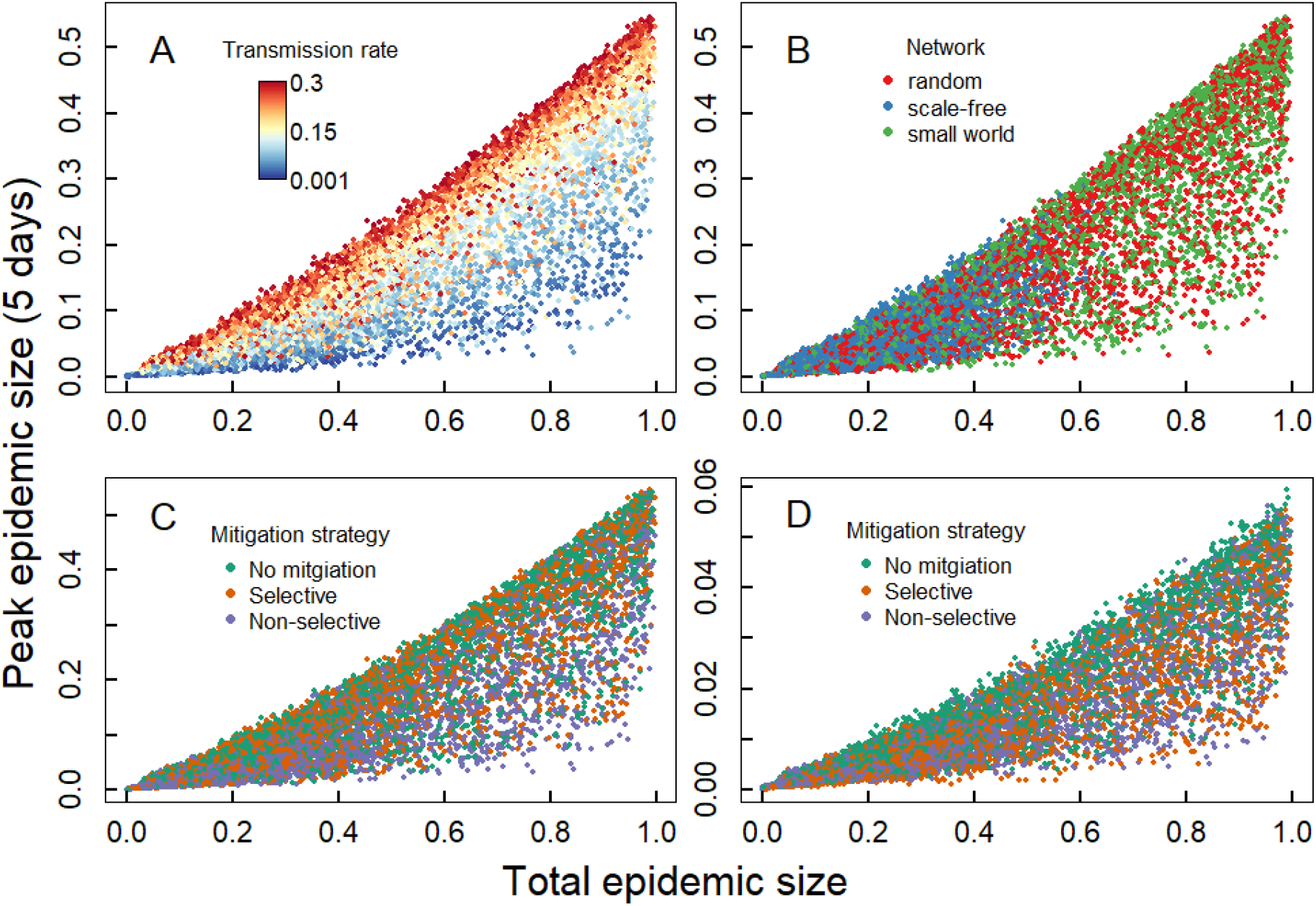
Relationship between total epidemic sizes and peak epidemic sizes for different transmission scenarios. Outcomes of different transmission scenarios in terms of total epidemic size (proportion of individual infected) and respective peak epidemic size (proportion of individuals infected in 5 days with most new infections). The different panels A-C show the same data for total peak epidemic sizes, plotted in different colour to show patterns in the data in relation to transmission rate (A), network type (B), and mitigation strategy underlying the modelled data. Panel D shows the peak epidemic sizes for high risk individuals only.

## Discussion

Our results highlight that strict short-term disease containment actions designed to halt the rapid increase in cases during peaks of local COVID-19, may have idiosyncratic outcomes and do not necessarily guarantee optimal protection of global populations over larger spatiotemporal scales. The modelled variation in underlying transmission scenarios may well represent the diversity of conditions and uncertainty we encounter in real worlds, resulted in a considerable number of scenarios having smaller total epidemic sizes without any stricter short-term containment measures. Selective containment measures can, under some conditions, protect more individuals form infections than apparently more costly whole-population containment efforts.

The intriguing result that short-term mitigation is not necessarily beneficial appears counterintuitive at first glance. This result, however is in line with general expectations of how metapopulations dynamics explain disease persistence alongside local suppression and spatial dissemination among connected populations (*9, 14, 15*). If short-term intensive containment efforts designed to flatten the epidemic curves of COVID-19 result in more prolonged (‘flattened’) local epidemics (*4*), disease spread among different populations can be particularly facilitated in highly connected populations and unfold its adverse effects at larger scale. This effect helps to explain the surprisingly low benefit of temporary mitigation strategies, including whole population isolation measures over several weeks or months.

Crucially, we do want to emphasize that containment efforts are absolutely vital for protecting people from infection by newly emerging disease such as COVI-19 until global vaccinations campaigns are in place to contain any further spread. We contend that constantly lowering transmission rates and carefully circumventing those links in metapopulation networks most supportive for disease spread, can ultimately reduce epidemic size at metapopulation scale. Efficient mitigation strategies are urgently needed to contain the spread of SARS-CoV-2 and save lives, but temporarily restricted intensive mitigation strategies (e.g. strict short-term containment and mass quarantine) in response to overloaded health care systems cannot replace efficient long-term mitigation strategies such as rigorous physical distancing or revisiting contact structure at landscape scale according to our results. Our results suggest that the ease or entire uplifting of existing intensive short-term containment efforts, such as mass quarantine, can be dangerous and may result in unprecedented disease spread. At the same time, achieving the long-term escape of vulnerable individuals from infection requires reliable containment measures that can be applied over the entire course of pandemics or until global vaccination campaigns offer better solutions. This may include measures to lower disease transmission such as protective gear and restructuring professional and social contact networks to resemble scale-free rather than small worlds or random connections among people and populations as much as possible. The dramatic differences in our model outcomes along the gradient of parameter values explored highlights the importance of accounting for regional conditions in real-world decision making. Both local human settlement density and connectivity structure, along with travel and social behaviour, which together set the stage for disease spread, differ widely across the globe. Informed decision making for rewiring interpersonal contacts or measures aimed at lowering transmission rates thus require taking regional conditions and feasibility into account. This knowledge will help policy makers planning adequate strategies to prevent the overwhelming of health care systems worldwide, and, ultimately, to protect lives.

## Data Availability

This is a theoretical study, model described in the Supplementary material, see methods and references for justification of parameters

## Acknowledgments

We thank our families for support while conducting this research in home offices.

## Author contributions

K.W. conceived the study and model, K.W. and M.L. analysed the data and wrote the paper.

## Data and materials availability

All code, and materials used in the analysis are available as Supplementary Materials.

## Supplementary materials

Materials and Methods

Figs. S1 to S3

Tables S1 to S5

## References and Notes

1. World Health Organization, Coronavirus Disease 2019 (COVID-19): Situation Report – 88 (17 April 2020). https://www.who.int/docs/default-source/coronaviruse/situation-reports/, (accessed 17/04/2020).

2. F. Zhou et al., Clinical course and risk factors for mortality of adult inpatients with COVID-19 in Wuhan, China: a retrospective cohort study. The Lancet. 395, 1054–1062 (2020). doi:10.1016/S0140-6736(20)30566-3

3. R. M. Anderson, H. Heesterbeek, D. Klinkenberg, T. D. Hollingsworth, How will country-based mitigation measures influence the course of the COVID-19 epidemic? The Lancet. 395, 931–934 (2020). doi:10.1016/S0140-6736(20)30567-5

4. B. F. Maier, D. Brockmann, Effective containment explains subexponential growth in recent confirmed COVID-19 cases in China. Science. eabb4557 (2020). doi:10.1126/science.abb4557

5. P. Fine, K. Eames, D. L. Heymann, “Herd immunity’’: a rough guide. Clin. Infect. Dis. 52, 911–916 (2011). doi:10.1093/cid/cir007

6. L. Ferretti et al., Quantifying SARS-CoV-2 transmission suggests epidemic control with digital contact tracing. Science. eabb6936 (2020). doi:10.1126/science.abb6936

7. L. Danon, J. M. Read, T. A. House, M. C. Vernon, M. J. Keeling, Social encounter networks: characterizing Great Britain. Proceedings of the Royal Society B: Biological Sciences. 280, (2013). doi:10.1098/rspb.2013.1037

8. H. Heesterbeek et al., Modeling infectious disease dynamics in the complex landscape of global health. Science. 347, (2015). doi:10.1126/science.aaa4339

9. M. J. Keeling, C. A. Gilligan, Metapopulation dynamics of bubonic plague. Nature. 407, 903–906 (2000). doi:10.1038/35038073

10. R. E. Rowthorn, R. Laxminarayan, C. A. Gilligan, Optimal control of epidemics in metapopulations. Journal of The Royal Society Interface. 6, 1135–1144 (2009). doi:10.1098/rsif.2008.0402

11. M. J. Keeling, P. Rohani, Modeling Infectious Diseases in Humans and Animals (Princeton University Press, 2008).

12. J. Hellewell et al., Feasibility of controlling COVID-19 outbreaks by isolation of cases and contacts. The Lancet Global Health. 8, e488–e496 (2020). doi:10.1016/S2214-109X(20)30074-7

13. M. E. J. Newman, The structure and function of complex networks. Siam Review. 45, 167–256 (2003). doi:10.1137/s003614450342480

14. C. Viboud et al., Synchrony, waves, and spatial hierarchies in the spread of influenza. Science. 312, 447–451 (2006). doi:10.1126/science.1125237

15. V. Colizza, A. Vespignani, Epidemic modeling in metapopulation systems with heterogeneous coupling pattern: theory and simulations. Journal of Theoretical Biology. 251, 450–467 (2008). doi:/10.1016/j.jtbi.2007.11.028

16. L. Danon, T. A. House, J. M. Read, M. J. Keeling, Social encounter networks: collective properties and disease transmission. Journal of the Royal Society Interface. 9, 2826–2833 (2012). doi:10.1098/rsif.2012.0357

17. G. Csardi, T. Nepusz, The igraph software package for complex network research. InterJournal, Complex Systems. 1695, http://igraph.org (2006).

18. M. Stein, Large sample properties of simulations using latin hypercube sampling. Technometrics. 29, 143–151 (1981).

19. R Development Core Team. (R Foundation for Statistical Computing, Vienna, Austria, 2020). https://cran.r-project.org/

20. S. A. Lauer et al., The incubation period of coronavirus disease 2019 (COVID-19) from publicly reported confirmed cases: estimation and application. Annals of Internal Medicine. (2020). doi:10.7326/M20-0504

21. J. M. Read, J. R. Bridgen, D. A. Cummings, A. Ho, C. P. Jewell, Novel coronavirus 2019-nCoV: early estimation of epidemiological parameters and epidemic predictions. medRxiv, 2020.2001.2023.20018549 (2020).

